# Healthcare worker reported barriers and potential facilitators of acute lower respiratory infection care delivery for children at Mchinji District Hospital in Malawi

**DOI:** 10.1101/2025.09.11.25335598

**Authors:** Shubhada Hooli, Beatwil Zadutsa, Everlisto Phiri, Karina Hofstee, Nichole Davis, Eric D. McCollum, Charles Makwenda, Carina King

## Abstract

**Background:** We sought to understand barriers to high quality hospital based acute lower respiratory infection (ALRI) care at Mchinji District Hospital in Malawi.

**Methods:** In 2020 we conducted focus group discussions (FGDs) with clinical officers (COs) who provided direct clinical care following a half-day refresher course on pediatric ALRI case management. The underpinning research methodology of the FGDs was phenomenology, and they were analysed using thematic analysis.

**Results:** We recruited 16 COs to participate in 3 FGDs. Five themes emerged: lack of confidence in ALRI diagnosis and management, high clinical burden with understaffing, dysfunctional team dynamics, limited physical resources, and the recognition of the importance of vital sign measurements despite barriers to practice.

**Conclusions:** COs shared several barriers and potential interventions to improve child ALRI care delivery at Mchinji District Hospital. Some solutions were locally implementable with minimal to modest cost such as a program for continuing education, standard operating procedures during electricity outages, and posting of job aides. However, many of their suggestions require investments and commitment from the Malawian Ministry of Health to increase staffing capacity and improve the physical infrastructure and are therefore of undetermined feasibility.

**Key Messages:** *What is already known on this topic:* ALRIs remain a leading cause of child mortality in Malawi, despite significant gains from the national child lung health program. Little is known about healthcare workers’ perspectives on hospital-based barriers to ALRI care.

*What this study adds:* Clinical officers described their lived experience of barriers to ALRI care delivery, including limited confidence in ALRI management, dysfunctional team dynamics, and resource shortages. They also identified potential solutions such as refresher trainings, job aides, structured plans to manage power outages, and improved access to pulse oximetry.

*How this study might affect research, practice, or policy:* We present healthcare worker-identified interventions that are perceived as acceptable and implementable. These lessons can serve as a starting point for both locally developed and national-level interventions and solutions.

## INTRODUCTION

Globally, between 1990 and 2021, there was a 50% reduction in the incidence of acute lower respiratory infections (ALRIs) and a 70% decrease in associated mortality amongst children under 5 years of age [1]. Despite these great strides, in 2021 over 500,000 children under 5 still died from ALRIs and most of these deaths occurred in low and middle-income countries (LMICs), such as Malawi [2].

In Malawi under 5 mortality decreased by two thirds between 1990 and 2013 to a mortality rate of 71 deaths per 1000 livebirths, achieving Millenium Development Goal 4 [3]. Much of this progress was attributed to early adoption of public health interventions including vaccination programs and increased access to treatment for diarrheal diseases and ALRI. In 2000, the Malawi Ministry of Health implemented the Child Lung Health Programme [4], which included the introduction of national clinical ALRI (otherwise referred to as pneumonia) diagnosis and management guidelines. The case fatality rate of children hospitalized with ALRI decreased from 15.2% in 2001 [5] to 3.2% in 2014 [6]. Despite an overall reduction in the child ALRI case fatality rate, minimal declines were observed for specific high-risk sub-populations, such as malnourished children, that currently account for most deaths.

A study from Malawi reported that over 50% of in-hospital deaths occur in the first 48 hours of admission [5]. Early hospital deaths can be due to a variety of factors such delays in care seeking, lack of care accessibility, or challenges with facility pediatric emergency readiness. A 2016 study concluded from Mchinji district, Malawi reported that 86% of suspected ALRI deaths sought care once and 44% did so at least twice, suggesting deaths were not predominantly the result of not seeking care [7]. Though hospital mortality is often attributed to delays due to long distances from care [8], in rural Malawi, research suggests quality of care is likely more important [9,10,11]. However, it is unclear if the in-hospital mortality burden is due to poor adherence of caregivers to treatment guidance [12] including fear of oxygen therapy [13], quality of health worker care delivery [14], a lack of human and hospital resources for emergency and ALRI care [15], or, a combination.

To understand the barriers to delivering high quality emergency and hospital-based care for children with ALRI, input from healthcare workers in these settings is needed. Though studies in Malawi have reported barriers, challenges, and outcomes of the implementation of care delivery programs such as Emergency Triage Assessment and Treatment [16,17] at primary care facilities, few if any have reported healthcare worker perspectives on their competency and ability to deliver child ALRI care within hospital settings. We therefore aimed to understand healthcare worker perceived barriers to the delivery of high-quality care to children admitted to a district hospital in Malawi with ALRI and report their perceptions of factors which may improve care. Our goal is to generate recommendations for potential interventions to further reduce in-hospital child ALRI mortality in rural Malawi.

## METHODS

We conducted a qualitative study, using focus group discussions (FGDs) with Clinical Officers (COs) engaged in child ALRI care at Mchinji District Hospital in Malawi in February 2020, as part of a mobile health (mHealth) decision support tool pilot study. The study was conducted and reported according to recommendations outlined in the Consolidated Criteria for Reporting Qualitative Research (COREQ) [18]. Neither patients, their parents/guardians/caregivers, nor the public were involved in the design of the research.

### Setting

Mchinji District Hospital (MDH) is in central Malawi near the Zambian border, with a catchment of approximately 500,000 people. In Malawi COs, a cadre of trained clinicians with three years of schooling, provide the majority of hospital based care [19]. At MDH COs are the primary providers for patients, leading their evaluation and management. They provide outpatient, emergency, and hospital based care. During after-hours one CO staffs the entire hospital with the support of nurses. Patients are generally evaluated by COs once daily with vital signs collected at that time. COs and nurses will periodically, and non-systematically, re-check patients they identified as high risk. At the time of the study there was one supervising physician at MDH who did not have subspecialty training in pediatrics. The paediatric ward at MDH had 50 beds, of which two were designed as high-dependency beds in a side-room next to a nursing station, with oxygen available primarily through a concentrator with a flow-splitter.

### Design

We conducted three FGDs following a half-day refresher course on child ALRI care with COs, located off hospital grounds. The purpose of the training (developed by BZ, EP, ND, and SH) was to remind participants of the Malawi Ministry of Health guidelines to ensure everyone was aware of what care was meant to be provided for children with ALRI, and help participants reflect on their current practice. The underpinning research methodology of the FGD was phenomenology so as to capture the lived experience of COs and minimize desirability bias.[20]

### Participants

Due to hospital staffing needs, nurses were not available to participate in the study. At the time the District Medical Officer was the only physician providing child ALRI care at MDH. Because of these limitations we decided to focus on only COs working at MDH were eligible to participate in the study. We used convenience sampling due to logistical constraints and the need for readily accessible participants with relevant experience. In the afternoon, following the training workshop, participants were approached for consent by study staff (BZ and EP). Workshop participants who provided verbal consent were then led to a separate room where FGDs occurred.

### Data Collection

FGDs were led by a male Malawian researcher (BZ) from the Parent and Child Health Initiative (PACHI), based in Mchinji district, and who was formerly a CO working on a paediatric ward. BZ was known professionally by the participants. He has led multiple qualitative studies focused on topics relevant to child ALRI care [16,21,22]. PACHI is a Malawian research nonprofit organization and had multiple programs and studies in the community. As these FGDs were coupled with a ALRI refresher course the participants were aware that we were interested in understanding barriers to child ALRI care. Only participants and researchers (BZ, SH, EP) were present during the FGDs. Both SH and EP, at times, asked clarifying questions.

The FGDs followed a structured guide (Supplement 1), which was not piloted prior to data collection. The central theme of questions was focused on CO perceived barriers to providing high quality child ALRI care at MDH across the continuum of outpatient, emergency, and inpatient care. FGDs lasted up to 1 hour, were audio recorded and then transcribed, and translated to English where necessary, by BZ and EP together. Participants were instructed at the start to use their preferred language, either English or Chichewa, with both BZ and EP fluent in both. Field notes were not recorded; there were no repeat interviews and participants did not have the opportunity to review transcripts for comment or correction.

### Analysis

The FGDs were analysed using thematic analysis [23]. Two members of the research team (SH and KH), both physicians with clinical experience in Malawi, independently coded transcripts. SH initially reviewed transcripts inductively to develop the initial coding tree. SH first noted subthemes and then categorized these into overarching themes. Then, utilizing the coding tree developed by SH, KH deductively coded the transcript and revised the initial coding tree. Finally, SH reviewed the transcripts and recoded them with the finalized coding tree. Data saturation was achieved after analyzing the final FGD. These data were then reviewed by members of the study team (BZ and CK) to ensure interpretation agreement. Data was managed in Microsoft Excel. All authors reviewed the codes, coding tree, and transcripts to assure consistency between the data and the findings. Participants were not given an opportunity to provide feedback.

### Ethics Statement

This study was approved by the Malawian National Health Sciences Research Committee under Protocol # 18/07/2098 and the Institutional Review Board of Baylor College of Medicine (H-43312). All participants provided informed verbal consent for both taking part in the FGD and the audio recording.

## RESULTS

We recruited a total of 16 participants (5-6 for each FGD), with all participants approached agreeing to participate. All participants were COs with experience providing child ALRI care at MDH. We identified 5 major themes with subthemes that reflect HCW perceived barriers to the delivery of high-quality child ALRI care at MDH (Figure 1). These themes reflect HCW confidence in their ability to manage child ALRI, a high clinical burden with relative scarcity of trained staff, limited physical resources at MDH, and barriers to vital sign measurement.

### COs lack confidence in ALRI diagnosis and management

Though at the time there had been no recent changes to ALRI guidelines in Malawi, COs expressed uncertainty in their clinical skills and knowledge of diagnosis and management, and that of their colleagues. Subthemes emerged expressing a desire to improve while also providing suggestions on how to maintain knowledge.

#### Desired skills and knowledge

Participants reported concerns about gaps in skills and knowledge to accurately diagnose ALRI. Many COs expressed a lack of confidence in their colleagues’ ability to diagnose and manage ALRI. They highlighted the need for training to build competence amongst COs.

> *“In diagnosing children with pneumonia the first problem we are facing is the skills. I would say most of us are incompetent in diagnosing pneumonia and in management “-Participant(P):5, Focus Group Discussion (FGD):3*

Many participants reported they had not had an ALRI refresher training since schooling. As the FGDs occurred directly after an ALRI training workshop, some COs expressed concerns that there were recommended medications that they had just been oriented on and were unfamiliar with. Others thought the guidelines provided in the refresher training was new.

> *“other challenges can be lack of update for new knowledge we can have new medicines which come for pneumonia but we cannot be updated so it has become more difficult to treat children using the drug which you are not familiar with because you are not updated on them”-P:4, FGD:2*

> *“some of these are new guidelines of which we don’t know we just learnt here, so I think it will be very crucial if we may introduce the new guidelines earlier than later because this maybe of beneficial to the health provider and the patients” - P:1, FGD:3*

#### Job Aides

CO emphasized the importance of readily accessible job aides. Easily referenceable guidance, such as process maps, were suggested. Multiple participants believed availability of these aides alone could improve child ALRI care. As one participant stated:

> *“ the first thing is we should have at least always the job aids that can assist us in diagnosing the pneumonias”-P:1, FGD:1*

> Another highlighted the need for visual aids, saying, “*we need these algorithms to be pasted everywhere we see patients, these algorithms will help us to remember, sometimes we forget”-P:4, FGD:1*

### High caseloads coupled with an inadequate number of trained staff result in burnout driven by futility

Healthcare workers described scenarios where there were more patients than could be possibly managed by trained staff. However, COs also acknowledged their colleagues are working the best they can under very difficult conditions. There were recurrent statements alluding to burnout and futility.

#### Inadequate Number of Trained Staff Relative to Patient Volume

Staff shortages are a major issue, leading to increased workloads for healthcare workers, and feelings of being overwhelmed. Participants reported this often results in rushed assessments and missed diagnoses, which may negatively impact patient outcomes. COs expressed a need for more trained staff as the current workforce is insufficient, and the high patient volumes further exacerbate the problem. COs felt pressure to assess patients as quickly as possible, making it difficult to provide the minimum standard of care. Their focus was on ensuring they see every patient in a timely manner.

> *“you end up being alone at room 1 [emergency triage area] so you can see that even when it is an emergency … [another] patient is coming in. You cannot be attending both of them at once [and] no one … is helping” -P:2, FGD:2*

> *“because of workload some might admit a patient … without checking the respiratory rate” -P:3, FGD:2*

> *“sometimes you are likely to miss other necessity right other important parameters for us to diagnose what pneumonia because you are under pressure and because of the queue so you just fast fast to let the patients go” -P:4, FGD:1*

#### Symptoms of Burnout/Futility

Though overall highly engaged participants expressed feelings of futility and burnout. There were expressions of feeling unsupported and isolated. At times they described instances managing critically ill children where they were the only clinician providing care.

> *“Sometimes we run with the patient in the corridors of the hospital with the patient looking for help so that your friends should assist you when you are doing other things, and another one doing other things, it’s difficult for one person to assist to start the medication on the admission room according if the child is critically sick” -P:2, FGD:1*

During one of the FGDs participants asked our research organization to help. They proposed ways we could fund additional staff and explicitly stated our focus should be on ways to supplement the care provided at MDH. These suggestions highlight feelings of futility working within the existing system.

> *“ PACHI you have to consider on addition of health workers more especially clinicians maybe on contract ”-P:1, FGD:1*

### The pressure of the system led to dysfunctional team dynamics and unapproved task shifting

There was a general sense of lack of support staff to aid the COs in care delivery. Some reflected skepticism about the work ethic of different support staff, though others acknowledged that they too were stretched to their limits. In response, participants reported untrained non-clinical hospital staff, such as housekeepers and janitors, volunteered to fill the gaps.

#### Cynicism about nursing work ethic

Participants reported nurses are supposed to measure vital signs of patients. Some reported concerns the nurses may not execute the tasks which they’re asked to do.

> *“the admission area you have a nurse but you can hardly see her maybe checking vital signs or doing whatever you become surprised that we are two here but what is my friend doing so sometimes it is just ..just an attitude of somebody” -P:1, FGD:1*

As the discussion evolved however, some COs acknowledged the nurses were also stretched quite thin given the few numbers and the high patient volumes.

> *“there are others nurses who try their best to take vitals especially in HDU where they see patients they do try their best but lack of resources” -P:3, FGD:1*

> *“it might be two nurses on duty in a ward the whole pediatric ward and it becomes difficult for them to help with vital signs so you have to do it yourself” - P:5, FGD:3*

#### Unsanctioned task shifting by lay staff

COs gave examples of non-clinical hospital workers providing clinical care. Often these workers did not have any formal training to measure vital signs, though some of the COs stated that they had taken the initiative to train specific individuals.

> *“In my mind I know some of the maids who do their job rightly or nicely to the patient the way they assess them, triaging them and some of them I trust them that they can take vital signs properly like they can touch them to assess for fever they can telling you this one can have high fever” -P:1, FGD:2*

Others expressed skepticism of unsanctioned task shifting. Though these workers might measure vital signs, they were described by some as being frequently inaccurate.

> *“Some patients attendants they do not know how to take for example the BP … he can of course give us the vital signs … if you go back and check the vital signs you see them very different from what you [obtain]” -P:6, FGD:2*

### Physical resource challenges negatively impact the quality of care

COs described multiple challenges with resources, ranging from a lack of basic supplies to infrastructure barriers such as a long distance between the emergency care area and the resuscitation room.

#### Lack of Basic Supplies

Essential medical equipment, including pulse oximeters, were in short supply. This prevented COs from diagnosing and treating ALRI effectively. It was especially notable in the resuscitation room where participants reported not having access to supplemental oxygen. Some reported that these resources were being shared amongst different areas of the hospital making them difficult to find when needed.

> *“we use it [pulse oximeter] when if its available but most of the time it’s not available actually, I should say we don’t have them” -P:4, FGD:3*

Some participants reported medical devices being stolen from the hospital. Though this was a widely held belief amongst the participants, the perception was that no one was following up on these concerns. Others acknowledged multiple areas of the hospital needed to use the same equipment and they believed the missing equipment was probably just misplaced.

> *“other tools are being stolen … because we don’t have tight security like in room 1 … some of the people that come and pick … equipment that we use for the patients [are] not … stealing … [its] because of lack of equipment in different wards” -P:2, FGD:2*

#### Infrastructural Barriers

The hospital infrastructure itself posed challenges. Issues such as the proximity of the emergency evaluation room where outpatients are sent before admission to the hospital ward, frequent power outages, lack of backup generators, and inadequate mosquito nets contribute to a suboptimal care environment. Some participants believed these problems caused patients to worsen or acquire new conditions (such as malaria) and resultantly increased mortality rates.

> *“We have no specific room for the very sick* patients *which may benefit from O_2_ therapy … there are 2 or 3 patients at the same bed using maybe one … oxygen concentrator” -P:6, FGD:2*

> *“ when the electricity is off those that are oxygen dependent, we lose them immediately”-P:5, FGD:1*

> *“many children start having other conditions for example malaria because the room is not protected from mosquitos we do not have mosquito barriers in the windows so our children are being bitten by the mosquitos and they get malaria while in the ward”-P:4, FGD:2*

### COs value vital sign measurements to guide ALRI care, but systemic barriers limit their measurement

COs understood that respiratory rate and oxygen saturation are cornerstones of ALRI diagnosis and severity assessment.

#### Recognition of high risk cases

Respondents seem to recognize particularly high-risk cases and the importance of measuring pulse oximetry and respiratory rate as part of clinical care. However, there are challenges to consistently performing these measurements due to workload and resource constraints.

> *“to spend another minute waiting for this child to take the respiratory rate it’s like you are worsening the condition rather rushing the patient to the ward”-P:6, FGD:2*

Medical equipment problems: There was a sheer scarcity of medical equipment and reports that, at times, when available they were not functional.

> *“lack of working tools as like pulse oximeter and resuscitation materials like O2 cylinders and the likes*” *-P:1, FGD:2*

## DISCUSSION

We explored CO perceptions of barriers and facilitators to providing high quality care for children with ALRI at Mchinji District Hospital in Malawi. Participants emphasized systems level issues that negatively impact their ability to diagnose ALRI and provide guideline-based care. These issues were multifactorial and included too few trained clinical staff, lack of equipment, and high patient volumes. These long-term issues impacted healthcare workers’ capacity and were linked to expressions of burnout and demotivation. Despite these challenges participants provided concrete and actionable suggestions to improve patient care, ranging from improved visibility of job aides, continuing professional development and education, to increased pulse oximeter access, changes to hospital layout, and larger scale programs that would require national investments.

Participants believed continuing education and integration of high visibility job aides alone could improve the quality of ALRI care delivery. Lack of job aides at healthcare facilities has been reported elsewhere [24], but it is difficult to discern if posting of job aides alone improves care as its generally part of a larger package of interventions. Ongoing continuing education is a potential strategy to improve child ALRI care, though previous studies of these interventions have reported mixed results. As an alternative to formal training sessions, during the COVID-19 pandemic, some programs developed e-learning systems. In Ghana HCWs were receptive to an e-learning and hybrid in person continuing professional development program, though notable limitations included data pricing and internet connectivity [25]. None the less, existing e-learning systems could be leveraged to develop hybrid continuing education programs with an in-person component or mentorship via text messaging.

COs recognized the important role pulse oximetry plays in risk stratification of child ALRI cases. Pulse oximeter use in clinical care is critical as hypoxemia is highly prevalent in children with ALRI, with a 2022 meta-analysis reporting approximately 23% of children with ALRI who were evaluated in LMIC clinics were hypoxemic [26]. Despite the WHO’s recommendation to use pulse oximetry, when it is available, to assess hypoxemia and clear evidence that pulse oximeters are essential medical devices,[27] many outpatient facilities and hospitals in LMICs do not have or use pulse oximeters in routine care [25, 27–32]. Pulse oximetry use may have spillover effects beyond correct administration of medical oxygen. In Malawi a study which re-trained COs in IMCI did not change antibiotic prescribing practices for child ALRI, but it improved provider confidence and ALRI classification. When this training was integrated with distribution of pulse oximeters antibiotic prescribing frequency decreased by 50% in comparison with controls, suggesting pulse oximetry served as an objective tool to assure the clinician it was safe to not prescribe antibiotics [28]. However, a scoping review reported pulse oximeter training alone does not increase use [29]. To successfully implement pulse oximetry there must be both pre-service and in-service training, coupled with supervision and mentorship [30].

Other suggested interventions which would require a greater investment at the hospital level included the reconfiguration of patient care spaces and development of a formal process to manage electricity outages. Few African studies have explored the impact of the built hospital environment on patient outcomes, though there are numerous in high-income settings [31], including subspecialty-driven guidelines for intensive care units [32]. A pediatric outpatient clinic in urban Malawi reported a 40% decrease in hospital mortality when changes were made to the physical space as part of a larger intervention which included increased staff, integration of patient flow processes including triage, and increased clinician training [33]. To understand the potential benefits of infrastructure improvements it would be helpful to quantify the problem such as the frequency of new malaria diagnosis after admission (reflective of the lack of mosquito nets) or inability to use available oxygen due to physical space – such as bed distance from oxygen sources. Stable electricity could also improve outcomes for patients. Malawi has frequent electricity outages and less than one third of health facilities have a source for back-up energy [34]. Those with back-up power should have standard operating procedures on outage management, including assigned responsibility for switching power sources if this is not automated. Specifically, inappropriate generator use can cause the fatal, difficult to recognize complication of carbon monoxide poisoning. When electricity is available at times it is unstable resulting in high or low voltage delivery (brown out) which degrades equipment. In the central region of Malawi, a hospital installed a solar power system to overcome these barriers, however faced some challenges with lack of local capacity to manage the system over time and the need for battery storage [35]. However local capacity to manage solar systems can be built, and has been shown to impact ALRI mortality. In Uganda solar-powered oxygen delivery was feasible and in a randomized controlled trial resulted in a relative mortality risk reduction of 48.7% with an estimated cost-effectiveness of $25 per Disability-Adjusted-Life Year (DALY) saved when used for child ALRI treatment [36]. A process to manage electricity outages at MDH would be helpful, but its impact on outcomes is uncertain without other interventions at the national government level.

Healthcare workers identified other solutions which require long term government investments to come to fruition such as improved medication stocking and expansion of clinical staff which could include task-shifting of vital sign measurements. A 2017 audit of 44 facilities in two districts in Malawi reported poor availability of pediatric formulations of essential medications. Namely amoxicillin suspension, the first line treatment for child ALRI, was only available in 33.3% of health facilities. Availability of cotrimoxazole, another antibiotic used to treat ALRI, was only 16.7% [37]. Health systems strengthening focused on improved consumable good availability coupled with increased staffing are together necessary to improve population health [38]. Despite ongoing efforts by the MOH to increase the number of healthcare workers, Malawi is not on track to meet the WHO standard of 4.45 health workers per 1000 people by 2040 [39]. As there isn’t the capacity to increase the healthcare workforce at Mchinji District Hospital immediately we anticipate, like in other settings, task shifting will continue. In Malawi, task shifting vital sign measurement and documentation can be effective, sustained, and improve outcomes [40,41]. It’s unclear if vital sign collection workers who have been informally trained – such as hospital cleaners trained by individual clinical officers – could be effective or potentially lead to harm.

Understandably, perhaps in response to uncontrollable factors, our findings suggested an undertone of CO burnout. Burnout amongst various cadres of Malawian HCWs across different disciplines is well documented before and after the COVID-19 pandemic [42,43]. Malawian clinicians with symptoms of burnout delivering HIV care more frequently self-reported suboptimal care provision in a study of clinicians working at 89 public health facilities in 2015 [44]. In a sub-sample of participants job-place level factors associated with burnout included an unsupportive supervisor and poor team dynamics [43]. Given the corrosive nature of burnout further compiling on the challenges of a resource constrained health system it can be daunting to address. A randomized control trial of an interactive chatbot reported short term outcomes of a modest reduction in behavioral health and burnout symptoms in Malawi CHWs, suggesting there may be implementable tools to reduce clinician burnout [45].

We had several limitations in our study. Notably we did not document the training, age, or length of employment of participants. Understanding participant experience and duration of employment, particularly in the public sector would add more context to our interpretation of the FGDs. Additionally, we did not engage other cadres of healthcare workers such as nurses, management staff, or untrained workers conducting task shifting, which would have allowed us to triangulate differing perspectives. To do so, during work hours, could have negatively impacted clinical care due to staffing shortages. Especially given the themes around task shifting and team dynamics, these other perspectives would be valuable to explore. Nonverbal cues may have been missed as we did not take field notes. Additionally, there is a risk for social desirability and researcher bias. Finally, participants were not given an opportunity to review the transcripts to assure they adequately reflected what they were communicating.

## CONCLUSIONS

In summary COs at Mchinji District Hospital shared several notable barriers and potential interventions to improve child ALRI care delivery at their hospital. A few of the proposed solutions are locally implementable with minimal to modest cost such as a hospital developed program for continuing education, standard operating procedures during electricity outages, or job aide availability. However, many of their suggestions require investments and commitments from the Ministry of Health to increase staffing capacity and improve the physical infrastructure of the hospital and are therefore of unexplored feasibility. Themes of burnout emerged which should be further explored locally to understand if there are contextually appropriate and feasible interventions. Despite these challenges clinicians remained hopeful and committed to serving their patients. Future directions of research should explore the impact of continuing education programs and informal vital sign assistants on clinical care delivery.

## Data Availability

Data cannot be shared publicly as though it was de-identified, if transcripts are scrutinized individuals could potentially be identified as their place of employment and time period was described. Data is available from the corresponding author upon request to researchers who meet the criteria for access to confidential data.

## AUTHOR CONTRIBUTIONS

Conceptualization: SH, BZ, EP, CM, EDM, CK; Data Curation: BZ, EP; Formal Analysis: SH, BZ, KH, CK; Funding Acquisition: SH; Investigation: SH, BZ, EP, CM, CK; Methodology: SH, BZ, EP, ND, CM, EDM, CK; Project Administration: SH, BZ, EP; Resources: SH, ND; Supervision: SH, BZ, CM, CK; Visualization: SH, KH; Writing – Original Draft Preparation: SH, CK; Writing – Review & Editing: SH, BZ, EP, KH, ND, CM, EDM, CK

## ACKNOWLEDGEMENTS

We express our gratitude to the clinical officers who participated in this study. In preparing this manuscript Grammarly was used to improve readability and its recommendations were subsequently reviewed and edited by the authors.

